# Flexibility of brain dynamics predicts clinical impairment in Amyotrophic Lateral Sclerosis

**DOI:** 10.1101/2022.02.07.22270581

**Authors:** Arianna Polverino, Emahnuel Troisi Lopez, Roberta Minino, Marianna Liparoti, Antonella Romano, Francesca Trojsi, Fabio Lucidi, Leonardo L. Gollo, Viktor Jirsa, Giuseppe Sorrentino, Pierpaolo Sorrentino

**Affiliations:** Institute of Diagnosis and Treatment Hermitage Capodimonte, 80131 Naples, Italy; Department of Motor and Wellness Sciences, University of Naples “Parthenope”, 80133 Naples, Italy; Department of Developmental and Social Psychology, University of Rome “La Sapienza”, 00185 Rome, Italy; Department of Advanced Medical and Surgical Sciences, University of Campania “Luigi Vanvitelli”, 81100 Naples, Italy; Turner Institute for Brain and Mental Health, School of Psychological Sciences, Monash University, 3800 Victoria, Australia; Institut de Neurosciences des Systèmes, Inserm, INS, Aix-Marseille University, 13005 Marseille, France; Institute of Applied Sciences and Intelligent Systems of National Research Council, 80078 Pozzuoli, Italy

**Keywords:** neuronal avalanches, functional repertoire, brain flexibility, brain dynamics, amyotrophic lateral sclerosis

## Abstract

Amyotrophic lateral sclerosis (ALS) is a multisystem disorder. This view is widely supported by clinical, molecular and neuroimaging evidence. As a consequence, predicting clinical features requires a comprehensive description of large-scale brain activity. Flexible dynamics is key to support complex adaptive responses. In health, brain activity reconfigures over time, involving different brain areas. Brain pathologies can induce more stereotyped dynamics, which, in turn, are linked to clinical impairment. Hence, based on recent evidence that brain functional networks become more connected as ALS progresses, we hypothesized that loss of flexible dynamics in ALS would predict their clinical condition.

To test this hypothesis, we quantified flexibility utilizing the “functional repertoire” (i.e. the number of unique patterns) expressed during the magnetoencephalography (MEG) recording, based on source-reconstructed signals. Specifically, 42 ALS patients and 42 healthy controls underwent MEG and MRI recordings. The activity of the brain areas was reconstructed in the classical frequency bands, and the functional repertoire was estimated to quantify spatio-temporal fluctuations of brain activity. In order to verify if the functional repertoire predicted disease severity, we built a multilinear model and validated it using a *k*-fold cross validation scheme.

The comparison between the two groups revealed that ALS patients showed more stereotyped brain dynamics (*P* < 0.05), with reduced size of the functional repertoire. The relationship between the size of the functional repertoire and the clinical scores in the ALS group was investigated using Spearman’s coefficient, showing significant correlations in both the delta and the theta frequency bands. In order to prove the robustness of our results, the *k*-fold cross validation model was used. We found that the functional repertoire significantly predicted both clinical staging (*P* < 0.001 and *P* < 0.01, in delta and theta bands, respectively) and impairment (*P* < 0.001, in both delta and theta bands).

In conclusion, our work shows that: 1) ALS pathology reduces the flexibility of brain dynamics; 2) sub-cortical regions play a key role in determining brain dynamics; 3) reduced brain flexibility predicts the stage of the disease as well as the severity of the symptoms. Based on these findings, our approach provides a non-invasive tool to quantify alterations in brain dynamics in ALS (and, possibly, other neurodegenerative diseases), thus opening new diagnostic opportunities as well as a framework to test disease-modifying interventions.

## Introduction

Amyotrophic Lateral Sclerosis (ALS) is caused by a combination of pathogenic processes, not limited to motor neurons, but rather involving the whole brain^1–3^. Neuroimaging studies confirm that ALS is a multisystem disorder characterized by alterations in motor and extra-motor brain regions^4–7^. Accordingly, despite the main symptoms being associated to motor dysfunction, up to 50% of ALS patients develop cognitive and/or behavioral impairment, and about 13% develop the behavioral variant of Frontotemporal Dementia (bvFTD)^1,3,8–10^. A strong pathophysiological link between these two pathologies is confirmed by the presence of a hexanucleotide repeat mutation in the *C9orf72* gene^11–13^ and by TAR-DNA binding protein-43 (TDP-43) inclusions observed in both ALS and FTD^10,14^. The abnormal accumulation of TDP-43 in neurons and glial cells of multiple brain regions has been identified as one of the major culprits in both ALS and FTD with ubiquitinated inclusions^13,15,16^.

Accounting for symptoms induced by widespread neurodegeneration requires a precise description of the fine-tuned, large-scale interactions among brain regions^17^. In fact, the large-scale brain organization has been shown to undergo extensive changes in ALS^18^. In particular, the progression of the disease shifts the (time-averaged) functional brain networks toward hyper-connectedness^19^.

The temporal organization of the interactions among regions is rich and contains both oscillatory and bursty (also referred to as “scale-free”, 1/f activity, fast transients or neuronal avalanches) components^20^. Such bursty activations account for most of the (time-averaged) functional connectivity^21,22^. Furthermore, rapid peaks of activations have been linked to behavioral outcomes, underlying their physiological significance^23^. Hence, time-averaged functional connectivity might not be enough to account for complex symptoms, and the specific, time-resolved patterns of activations might instead be relevant. In fact, complex brain functions require both reliable communication among brain regions as well as the prompt ability to reconfigure their interactions^24,25^. From a physical perspective, it was posited that the brain operates in a near-critical regime, which is at the edge of a phase transition and optimizes computational functions^26^. This fact would account for many statistical properties that are observed in real data, and large-scale bursts of activations are interpreted as “neuronal avalanches”, which occur at multiple spatial and temporal scales^27^. Neuronal avalanches are tightly regulated and their spatial spreading pattern is constrained by the structural connectome^28^. In a healthy brain, neuronal avalanches constantly reconfigure over time, and recruit different groups of regions, resulting in the brain exploring a large number of patterns. The number of patterns, or size of the functional repertoire, provides a measure of brain flexibility, and is linked to healthy brain functioning. For example, a reduction of the functional repertoire relates to clinical impairment in Parkinson’s disease^29^. Hence, given the hyper-connected static functional topology observed in ALS^19^, we hypothesized that brain dynamics would be more stereotyped in ALS patients as compared to healthy controls. Furthermore, if complex brain functions require flexibility, and brain impairments derived from ALS could impair flexibility, a restriction of the functional repertoire should indicate functional impairment.

To estimate flexibility, we used source-reconstructed magnetoencephalographic (MEG) data, which have a time resolution of milliseconds. We filtered the signal in the classical frequency bands, and defined a *neuronal avalanche* as an event that begins when at least one brain region deviates from its baseline activity, and ends when all regions restore their typical level of activity (Fig. 1). Given a neuronal avalanche, we defined its corresponding *pattern* as the set of all the brain areas that were recruited. Finally, we defined the *functional repertoire* as the set of the unique patterns that happened over time (i.e. the part of the state-space that has been visited), and used its size as a proxy for the flexibility of the brain dynamics. We further hypothesized that the restriction of the functional repertoire would indicate worse clinical conditions. To test this claim, we built a model to predict the disease severity starting from the functional repertoire of ALS patients.

**Figure 1.**
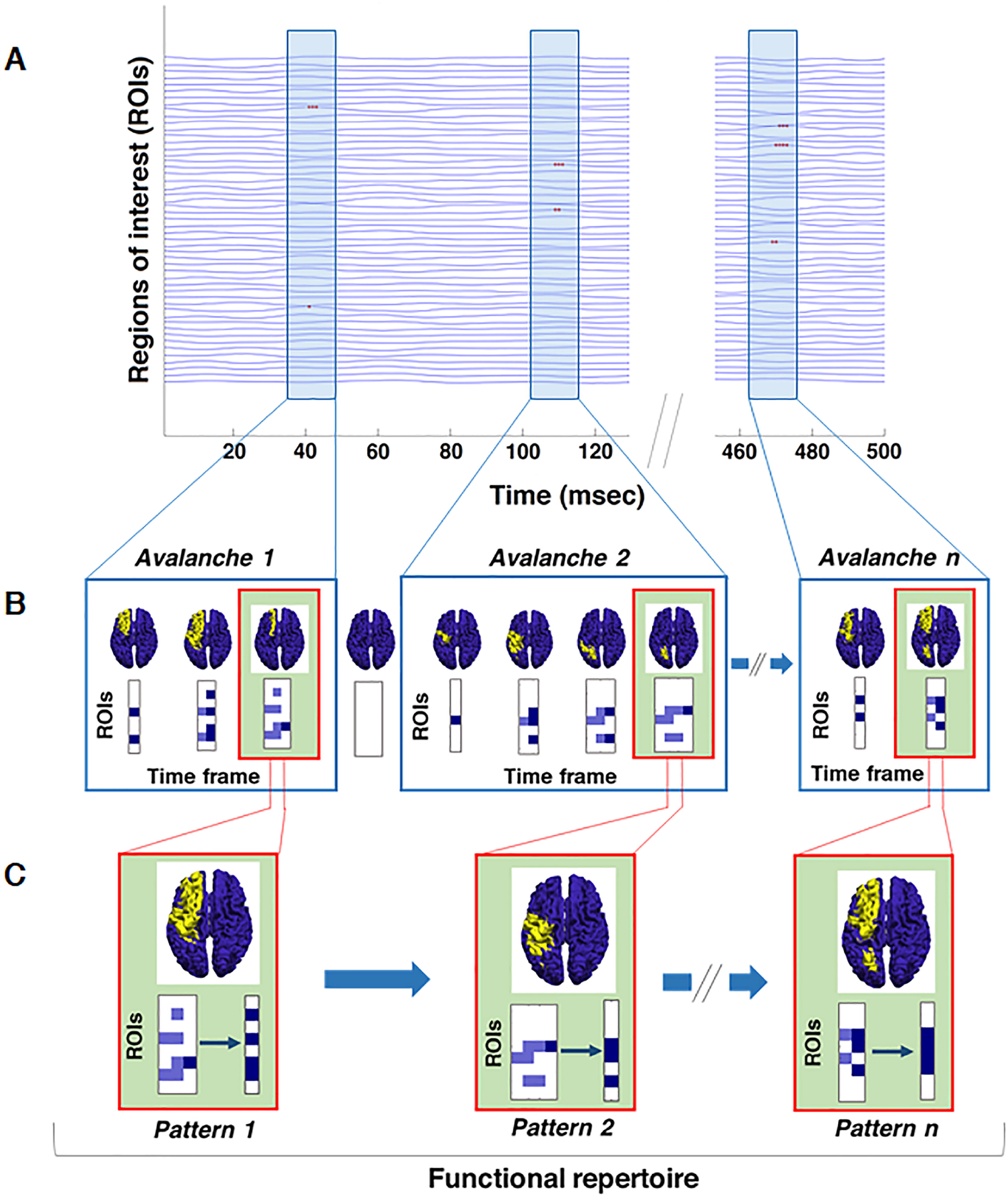
Schematic representation of neuronal avalanches and functional repertoire. (**A**) Source-reconstructed time series. Light blue rectangles represent the time lapses in which neuronal avalanches occur; red dots in the rectangles define activated brain regions (signal above the threshold) in a certain time interval (msec). (**B**) In the boxes, avalanche patterns of three different neuronal avalanches are illustrated (for simplicity, here avalanches consist of a maximum of 4 time frames). An avalanche is defined as an event that begins when at least one brain region deviates from its baseline activity (above threshold) and ends when all regions display a typical level of activity (below threshold). Given a neuronal avalanche, its corresponding pattern is the set of all the brain areas that were recruited at any time. The brains-plots for each time frame of an avalanche show the areas above (yellow) and below (blue) threshold. Each matrix represents an avalanche pattern: dark blue squares indicate the brain regions (ROIs) activated at a certain time frame, while the light blue ones are all the regions that have been activated until that moment. (**C**) In the green boxes, for each avalanche above, the brain-plot and the set of unique avalanche patterns are illustrated. Unique means that each avalanche pattern only counts once. The number of unique avalanche patterns defines the size of the functional repertoire and is used as a proxy for the flexibility of the brain dynamics.

## Materials and methods

### Cohort description

Forty-two ALS patients (32 males, 10 females; mean age ± SD, 64.81 ± 12.83) were recruited from the ALS Center of the First Division of Neurology of the University of Campania “Luigi Vanvitelli” (Naples, Italy). Patients were right-handed and native Italian speakers diagnosed with ALS according to the revised El-Escorial criteria of ALS^30^. None of the patients showed any mutation in the screened genes *SOD1, TARDBP, FUS/TLS*, and *C9ORF72*. Forty-two age-matched healthy controls (28 males, 14 females; mean age ± SD, 63.10 ± 10.46) were also included in the study.

For clinical assessment, we used the total Amyotrophic Lateral Sclerosis Functional Rating Scale-Revised (ALSFRS-R)^31^. To quantify disease staging we used the ALS clinical staging systems. Hence, patients were also classified according to both the King’s^32^ and the Milano-Torino Staging (MiToS)^33^ disease staging systems which are based on the appearance of sequential clinical milestones during ALS.

More clinical details and descriptive information about the cohort are reported in **Table 1**. The following inclusion criteria were used: (1) no use of drugs that could interfere with MEG signals; (2) no other major systemic, psychiatric or neurological diseases; and (3) no focal or diffuse brain damage at routine MRI. The study protocol was approved by the Local Ethics Committee (University of Campania “Luigi Vanvitelli”) with protocol number 591/2018, and all participants provided written informed consent in accordance with the Declaration of Helsinki.

**Table 1.** Demographic and clinical features of the cohort recruited for the study.

| Parameters | ALS patients (n = 42),<br>mean (± SD) | HC (n = 42),<br>mean (± SD) | P-values |
| --- | --- | --- | --- |
| **Demographic and clinical measures** |  |  |  |
| Age | 64.81 (± 12.83) | 63.10 (± 10.46) | 0.50 |
| Male/female | 32/10 | 28/14 | 0.0001*** |
| Education | 10.21 (± 4.61) | 12.38 (± 4.21) | 0.03* |
| Disease duration (months) | 49.21 (± 58.07) | | |
| ALSFRS-R | 36.08 (± 8.24) | | |

Significance *P*-values: \**P* < 0.05, \*\*\**P* < 0.001.
ALS = Amyotrophic Lateral Sclerosis; ALSFRS-R = Amyotrophic Lateral Sclerosis Functional Rating Scale-Revised; HC = healthy controls; SD = standard deviation.

### Brain network analysis

#### MRI acquisition

As previously described^34^, MRI images of 35 patients and 31 healthy controls were acquired on a 3T scanner equipped with an 8-channel parallel head coil (General Electric Healthcare, Milwaukee, WI). MR scans were acquired after the MEG recording or at least a month before. In particular, three dimensional T1-weighted images (Gradient-echo sequence Inversion Recovery prepared Fast Spoiled Gradient Recalled-echo, time repetition = 6988 ms, TI = 1100 ms, TE = 3.9 ms, flip angle = 10, voxel size = 1 × 1 × 1.2mm^3^) were acquired. The remaining participants (7 patients and 11 controls) did not complete the MRI because of the difficulty in lying down or because they refused to perform the MRI scan. A standard MRI template was used in this case.

#### MEG acquisition

The MEG system was developed at the Institute of Applied Sciences and Intelligent Systems “E. Caianiello” of the National Research Council, Pozzuoli, Naples^35^. MEG data were acquired with a 163-magnetometers system placed in a magnetically shielded room (AtB Biomag UG, Ulm, Germany) to reduce background noise. Data acquisition, pre-processing, and source reconstruction were as previously described^36^.

Briefly, before each acquisition, four reference positions (nasion, right, and left preauricular and apex) were digitalized on the subject’s head using Fastrak (Polhemus®). Electrocardiographic and electrooculographic signals were co-recorded^37^.

The brain activity of each subject was recorded in two segments, each 3.5 minutes long, during resting state with eyes closed. The instructions were delivered immediately before each recording via intercom. Data were acquired with a sampling frequency of 1024 Hz, and a 4th-order Butterworth IIR band-pass filter was then applied to remove components below 0.5 and above 48.0 Hz^38^. The filter was implemented offline using MatLab scripts within the Fieldtrip toolbox 201456^39^.

#### Data pre-processing

At this stage, the principal component analysis (PCA) was used to orthogonalize the sensors over the base of the reference sensor, as to remove environmental noise. Then, after visual selection of the data from an experienced operator, supervised independent component analysis (ICA) was used to remove physiological artifacts such as electrocardiogram and eye blinks (if present).

#### Source reconstruction

Source reconstruction was performed using a beamforming procedure implemented in the Fieldtrip toolbox^39^, similarly to Sorrentino and colleagues^19^. Firstly, the fiducial points of each participant were used to co-register the MEG data to the native subject-specific MRI. Subsequently, using the brain volume conduction model proposed by Nolte^40^, a Linearly Constrained Minimum Variance (LCMV) beamformer^41^ was used to reconstruct time series related to the centroids of 116 regions of interest (ROIs), derived from the Automated Anatomical Labeling (AAL) atlas^42,43^. Both the atlas and the MRI were aligned to the head coordinates. For each source, we projected the time series along the dipole direction that explains the most variance by means of singular value decomposition. We then considered the first 90 ROIs for further analysis, excluding the cerebellum given the low reliability of the reconstructed signal in this region^44^. The source-reconstructed signals were filtered in the classical frequency bands: delta (0.5 – 4.0 Hz), theta (4.0 - 8.0 Hz), alpha (8.0 - 13.0 Hz), beta (13.0 - 30.0 Hz) and gamma (30.0 - 48.0 Hz).

### Analysis of brain dynamics

#### Neuronal avalanches and branching parameter

To quantify spatio-temporal fluctuations of brain activity, we first estimated neuronal avalanches. As previously described^45,46^, an avalanche is defined as an event starting when an unexpected fluctuation of regional activity is present, and ending when all regions are inactive and back to normal activity. The number of events in all ROIs in an avalanche corresponds to its size. Note that the position of the brain regions in space are not considered, therefore two co-activated areas (in time) may even be located in different hemispheres.

Each of the 90 source-reconstructed signals were *z*-transformed. Subsequently, each time series was thresholded according to a cut-off of 3 standard deviations (i.e., *z* > |3|)^29^. To confirm that the results are not dependent upon the choice of a given threshold, we repeated the analyses setting the threshold to *z* > |2.5| and *z* > |3.5|.

To select a suitable bin length, we computed the branching ratio *σ*^47,48^ as follows: for each subject, for each time bin size, and for each avalanche, the geometrically averaged ratio of the number of events (activations) between the subsequent time bin and that in the current time bin was calculated as

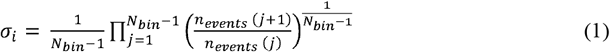

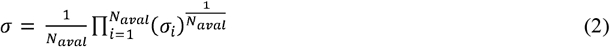

where *σ*_*i*_ is the branching parameter of the *i*-th avalanche in the dataset, *N*_*bin*_ is the total amount of bins in the *i*-th avalanche, *n*_*events*_ (*j*) is the total number of events active in the *j*-th bin, and *N*_*aval*_ is the total number of avalanches in each participant’s recording.

In branching processes, a branching ratio of *σ* = 1 indicates critical processes with activity that is highly variable and nearly sustained, *σ* < 1 indicates subcritical processes in which the activity quickly dies out, and *σ* > 1 indicates supercritical processes in which the activity increases as runaway excitation. The bin length equal to three samples yielded a critical process with *σ* = 1 hinting at the avalanches as occurring in the context of a dynamical regime near a phase transition. This means that each bin is obtained from three time-points of the binarized time-series. In order to confirm the robustness of our results, we investigated different time bins, ranging from 1 to 5. To equally compare the dynamics of brain activity among the subjects, we took into consideration the same duration for each participant time series (122.79 seconds). Segments of equal duration were randomly selected from the whole recording. For each avalanche, an *avalanche pattern* was defined as the set of all areas that were above threshold at some point during the avalanche.

#### Functional repertoire

For each participant we estimated the *functional repertoire*, defined as the number of unique avalanche patterns expressed during the recording^29^. *Unique* indicates that each avalanche pattern only counts once towards the size of the functional repertoire (i.e., it does not matter if a given avalanche pattern appears only once or multiple times, as only the number of different patterns contributes to the functional repertoire). A representation of avalanche patterns and functional repertoire is shown in **Fig. 1**.

#### Switching between states

A *switch* represents the exceeding of the threshold level, in either direction, and therefore occurs when an active region becomes inactive, and vice versa, between two consecutive time bins. The switch rate (number of switches over duration), averaged over areas, was computed for each participant.

#### Regional influence on avalanche patterns

At this stage, we split the total functional repertoire into two groups: patterns that occurred in both the clinical and control participants (“shared repertoire”), and patterns that were unique to either group (“group-specific repertoire”). Then, using the Kolmogorov-Smirnov test, we compared the distributions of brain regions occurrences between shared and group-specific repertoires, and performed permutation testing to identify which brain regions occurred significantly more in group-specific repertoire than in shared repertoire. We then tested if these occurrences were higher in the healthy or the patient group.

#### Multilinear model analysis

We then moved on to test the hypothesis that efficient brain dynamics is related to the correct functioning of the brain and, hence, the restriction of the functional repertoire related to clinical impairment. To test this hypothesis, we built a multilinear model to predict clinical measures and the stage of the disease based on demographics, clinical and the efficiency of brain dynamics (as measured by the size of the functional repertoire)^49^. Specifically, we considered the ALSFRS-R and the stage of the disease (King’s and MiToS clinical staging systems) as dependent variables, while age, education level, gender, disease duration and size of the functional repertoire were considered as predictors. Multicollinearity was assessed through variance inflation factor (VIF)^50,51^. In order to strengthen the reliability of our model, we performed the *k*-fold cross-validation, with *k* = 5^52^. Specifically, *k* iterations were performed to train our model and at each iteration the *k*^*th*^ subgroup was used as a test set.

The same design was also repeated using the leave-one-out cross validation (LOOCV). Expressly, we built *n* multilinear model (where *n* is the size of the sample included in the model), each time excluding a different subject from the model, and verifying the ability of the model to predict the clinical value of the excluded subject.

### Statistical analysis

To compare age and educational level between ALS patients and healthy controls we performed a T-test, while Chi-square was used for gender comparison. Permutation testing or Kolmogorov-Smirnov test was performed to compare patients and controls, as appropriate. For permutation testing, the data were permuted 10,000 times, and at each iteration the absolute value of the difference between the two groups was observed, building a null distribution of absolute differences. Finally, the empirical observed difference was rank-ordered against this distribution, yielding a significance value. The relationship between the size of the functional repertoire and clinical scores was investigated in the ALS group using Spearman’s correlation coefficient.

Results were corrected by False Discovery Rate (FDR), for both parameters and frequency bands, and the significance level was set at *P*-value < 0.05. The reported significances refer to the corrected ones as described. All statistical analyses were performed using custom scripts written in MatLab 2019a.

### Data availability

The code will be made available on GitHub upon acceptance. The data cannot be made available given its clinical nature. The data can be made available upon request to the first authors conditional to acceptance by the Ethical Committee.

## Results

### Functional repertoire, avalanche patterns and local dynamics

We used source-reconstructed resting-state MEG data acquired from a cohort of 42 ALS patients and 42 age-matched healthy controls in order to quantify the functional repertoire, which is the total number of unique avalanche patterns occurred in each individual. A comparison between the two groups revealed that ALS patients expressed a restricted functional repertoire, with a lower number of visited patterns. More specifically, we observed these results in both the delta (*P* = 0.046; **Fig. 2A**) and the theta (*P* = 0.046; **Fig. 2B**) frequency bands.

**Figure 2.**
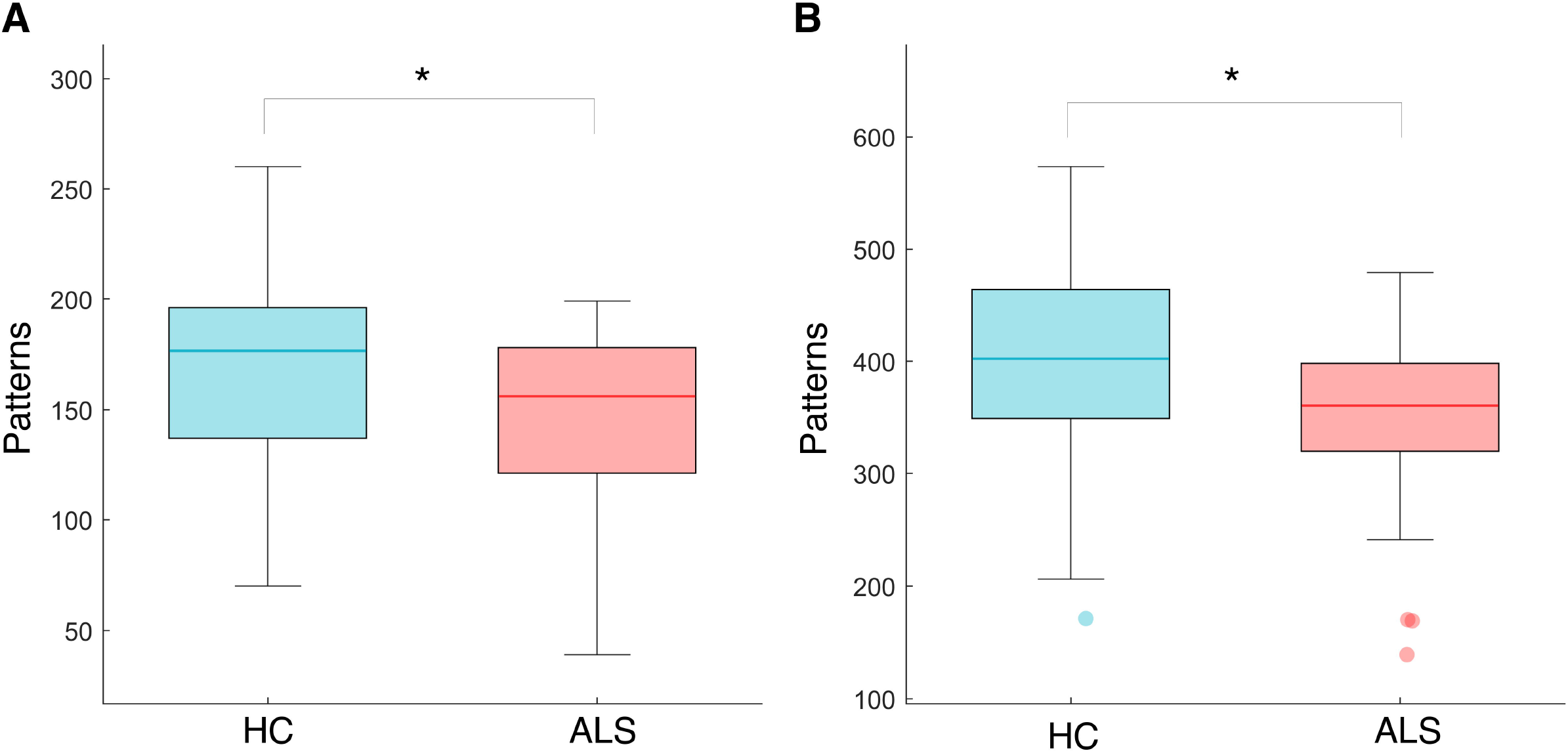
Comparison of the number of unique avalanche patterns. Box plots illustrating differences in the size of the functional repertoire in healthy controls (HC) and ALS patients (ALS), in delta (**A**) and theta (**B**) frequency bands. The central mark in the box indicates the median, the edges of the box the 25th and 75th percentiles and the whiskers extend to the 10th and 90th percentiles. The outliers are plotted individually using dots. Significance *P*-value: \**P* < 0.05. The figure was made using MatLab 2019a.

In order to prove the robustness of our results to specific choices of the avalanche threshold and bin length, we tested these variables across a moderate range of values and repeated the analyses. Specifically, we first used different binnings, ranging from 1 to 5. The results remained unchanged, with ALS patients displaying a restricted functional repertoire for all the binnings explored (for binning = 2, *P* = 0.009 (delta band) and *P* = 0.010 (theta band); for binning = 3, *P* = 0.010 (delta) and *P* = 0.012 (theta); for binning = 4, *P* = 0.010 (delta) and *P* = 0.015 (theta); for binning = 5, *P* = 0.011 (delta) and *P* = 0.019 (theta); see **Supplementary Fig. 1**). Furthermore, the avalanche threshold was modified, ranging from 2.5 to 3.5. For both cases the differences between the groups were confirmed (for *z* = 2.5, *P* = 0.028 in delta frequency band and *P* = 0.030 in theta frequency band, while for *z* = 3.5, *P* = 0.005 and *P* = 0.007 in delta and theta bands, respectively; see **Supplementary Fig. 2**).

We also evaluated how many times each active region became inactive, and vice versa (number of switches), finding no significant differences between the two groups (data not shown). These results confirm that brain dynamics is qualitatively altered in ALS patients, as compared to controls. In fact, the same number of switches means that the rate at which each region changes its status is similar in the two groups. Nonetheless, patients only visit a restricted number of patterns as compared to controls. Hence, the restriction of the functional repertoire is not due to different activation rates, but to a reduced number of combinations that the active regions produce in time.

Subsequently, we investigated the influence of specific regions on avalanche patterns. Firstly, we compared the distributions of brain regions occurrences between shared and group-specific patterns. The Kolmogorov-Smirnov test confirmed that the two distributions were significantly different (*P* < 0.001), meaning that there was an uneven involvement of brain regions in the avalanches of the two groups. Hence, we conducted a post-hoc analysis through a permutation test to identify which brain areas occurred more often in the group-specific patterns. The analysis highlighted several brain regions occurring significantly more in the group-specific repertoire than in shared repertoire (**Fig. 3**). In particular, in the delta frequency band, the right insula (*P* = 0.011), the right putamen (*P* = 0.023) and the pallidum bilaterally (*P* = 0.034 and *P* = 0.049, right and left, respectively) were more often involved in avalanches in the ALS group than in the controls (**Fig. 3A**). Similarly, in the theta band, the right Heschl’s gyrus (*P* = 0.025), the right putamen (*P* = 0.031), the right pallidum (*P* = 0.011) and the right thalamus (*P* = 0.045) occurred mainly in the ALS-specific repertoire (**Fig. 3B**).

**Figure 3.**
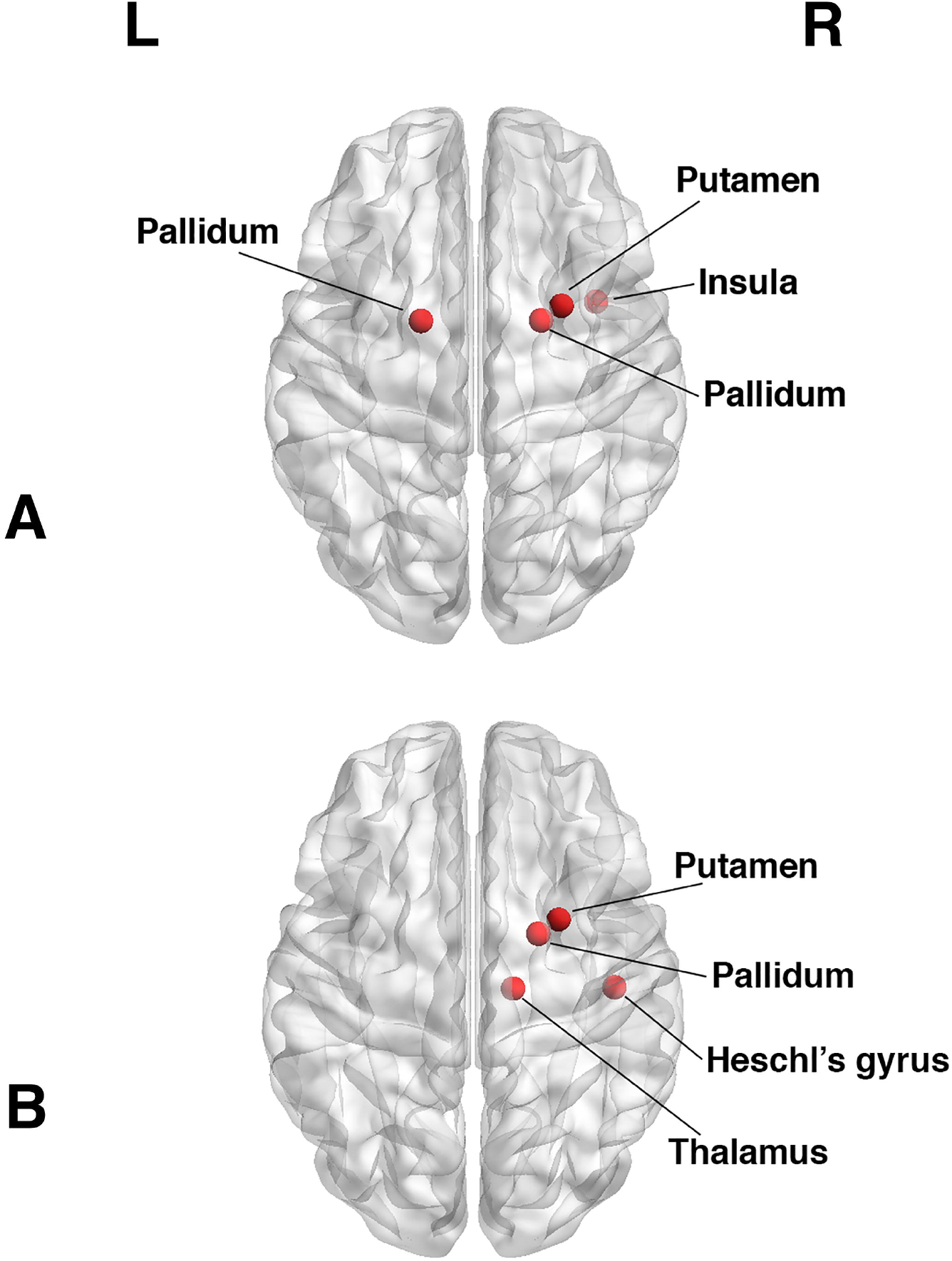
Mapping of brain regions occurring significantly more in the ALS-specific unique avalanche patterns. In particular, in the delta frequency band (**A**), the right insula (*P* = 0.011), the right putamen (*P* = 0.023) and the pallidum bilaterally (*P* = 0.034 and *P* = 0.049, right and left, respectively) are more often involved in avalanches in the ALS group than in the controls. Similarly, in the theta band (**B**), the right Heschl’s gyrus (*P* = 0.025), the right putamen (*P* = 0.031), the right pallidum (*P* = 0.011) and the right thalamus (*P* = 0.044) occur mainly in the ALS-specific repertoire. Significance *P*-value: *P* < 0.05. The image was made using MatLab 2019a, including BraiNetViewer v. 1.62.

### Multilinear model analysis

Subsequently, to understand the clinical significance of the restricted functional repertoire observed in ALS patients, we correlated the size of the functional repertoire to clinical measures such as disease duration, the ALSFRS-R, and both the King’s and the MiToS clinical staging systems.

We observed significant correlations between the number of avalanche patterns and clinical features in both the delta and the theta frequency bands. Particularly, in the delta band the number of distinct patterns correlates positively with the ALSFRS-R (*R* = 0.37, *P* = 0.019; see **Fig. 4A**), and negatively with both the King’s (*R* = -0.51, *P* = 0.002; see **Fig. 4B**) and the MiToS (*R* = -0.41, *P* = 0.015; see **Fig. 4C**) disease staging systems. These results can also be observed in the theta frequency band, where the number of avalanche patterns correlates positively with the ALSFRS-R (*R* = 0.40, *P* = 0.015; see **Fig. 5A**) and negatively with the King’s (*R* = -0.53, *P* = 0.002; see **Fig. 5B**) and the MiToS (*R* = -0.47, *P* = 0.005; see **Fig. 5C**) staging systems.

**Figure 4.**
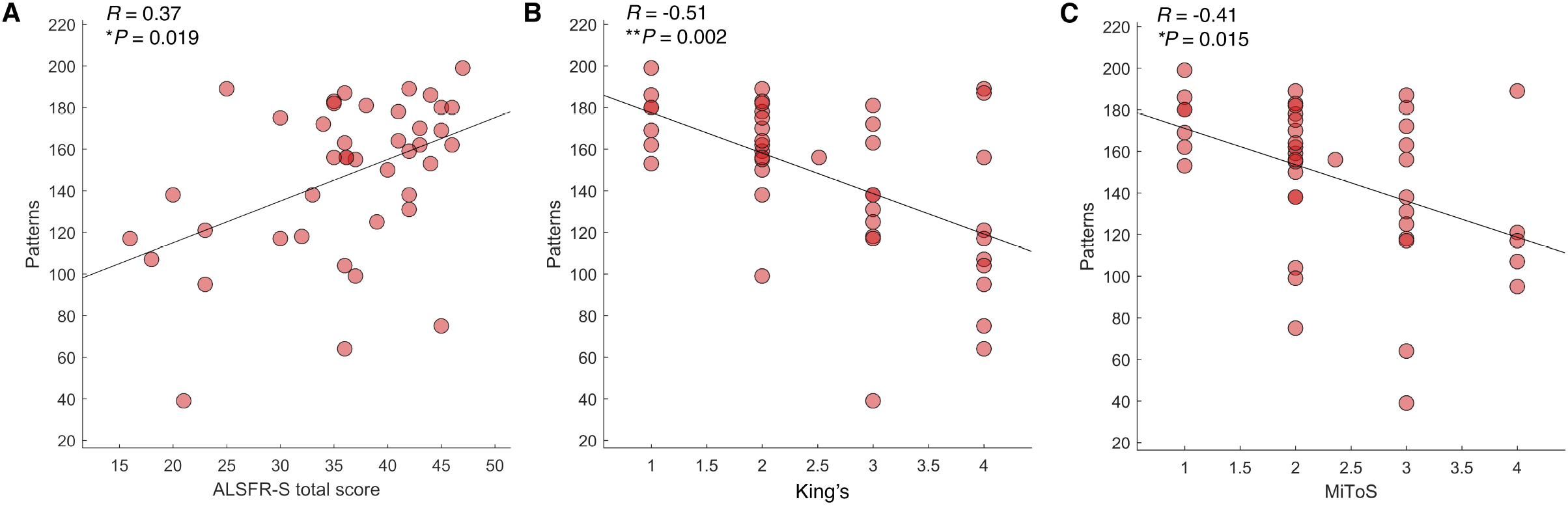
The relationship between brain dynamics and clinical features in the ALS group in the delta frequency band. (**A**) Positive correlation between the number of unique avalanche patterns and the ALSFRS-R (*R* = 0.37, *P* = 0.019); (**B**) negative correlation between the number of patterns and the King’s clinical staging system (*R* = -0.51, *P* = 0.002); (**C**) negative correlation between the size of the functional repertoire and the MiToS staging system (*R* = -0.41, *P* = 0.015). Spearman’s correlation coefficient was used and results were corrected by False Discovery Rate (FDR) correction. Significance *P*-values: \**P* < 0.05, \*\**P* < 0.01. The figure was made using MatLab 2019a.

**Figure 5.**
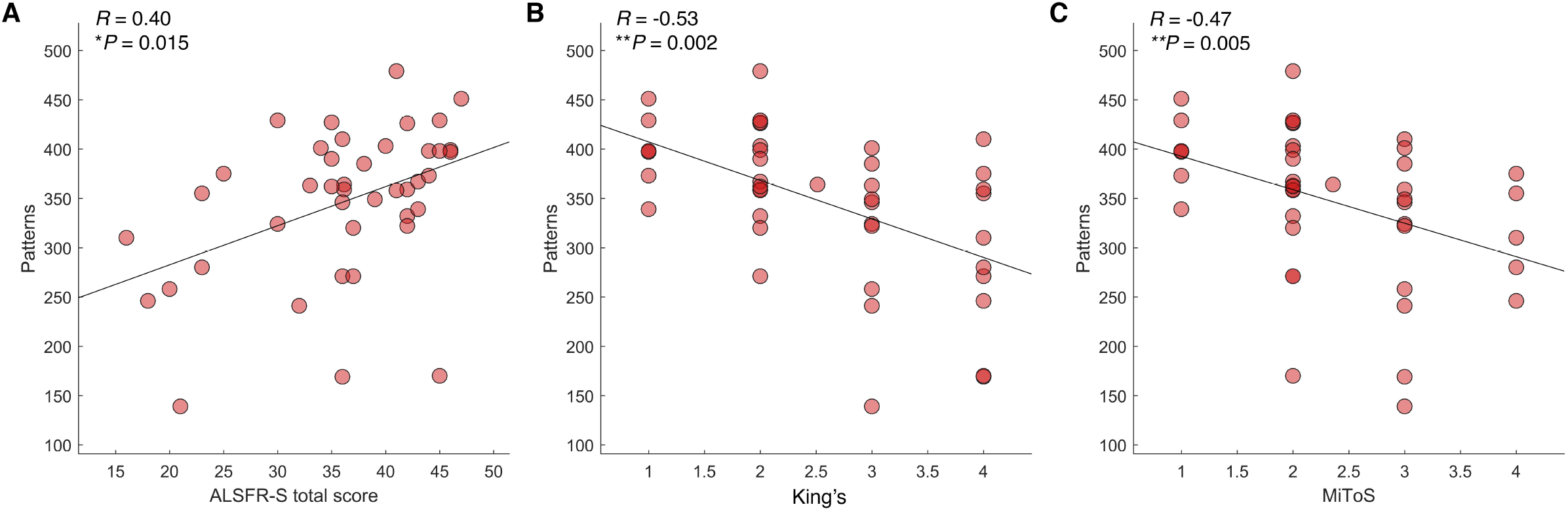
The relationship between brain dynamics and clinical features in the ALS group in the theta frequency band. (**A**) Positive correlation between the number of unique avalanche patterns and the ALSFRS-R (*R* = 0.40, *P* = 0.015); (**B**) negative correlation between the number of patterns and the King’s clinical staging system (*R* = -0.53, *P* = 0.002); (**C**) negative correlation between the size of the functional repertoire and the MiToS staging system (*R* = -0.47, *P* = 0.005). Spearman’s correlation coefficient was used and results were corrected by False Discovery Rate (FDR) correction. Significance *P*-values: \**P* < 0.05, \*\**P* < 0.01. The figure was made using MatLab 2019a.

Then, we used a multilinear model analysis with *k*-fold cross-validation to evaluate if demographics, clinical and brain dynamics features could predict the ALSFRS-R and the stage of the disease. We found that the size of the functional repertoire significantly improves the predictive capacity of the model in both the delta and the theta frequency bands (**Fig. 6** and **Fig. 7**).

**Figure 6.**
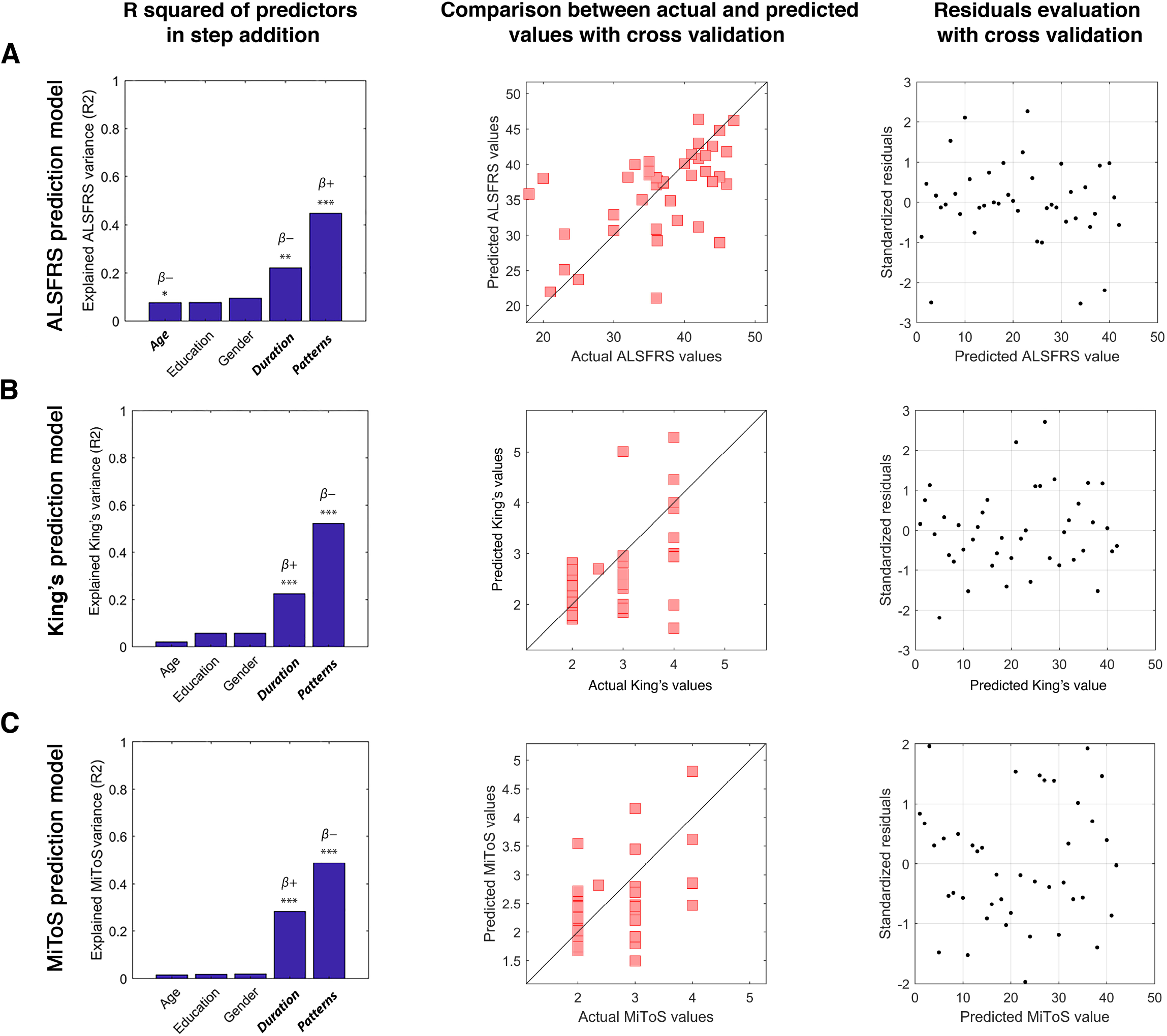
Multilinear model with *k*-fold cross-validation in the delta frequency band. Using as predictors age, education, gender, disease duration and number of patterns, the model predicts: (**A**) the ALSFRS-R (age: *P* = 0.036, *β* = -0.17; disease duration: *P* = 0.004, *β* = -0.06; number of patterns: *P* < 0.001, *β* = 0.11); (**B**) the King’s clinical staging system (disease duration: *P* < 0.001, *β* = 0.01; number of patterns: *P* < 0.001, *β* = -0.02); (**C**) the MiToS clinical staging system (disease duration: *P* < 0.001, *β* = 0.01; number of patterns: *P* < 0.001, *β* = -0.01). In the left panel of each row, the explained variance of the variable to be predicted as a function of the predictors is illustrated. Significant predictors are indicated in bold; positive and negative coefficients are illustrated with *β+* and *β-*, respectively; significance *P*-values: \**P* < 0.05, \*\**P* < 0.01, \*\*\**P* < 0.001. In the central panel of the rows, scatter plots of the comparison between actual and predicted values are represented. The standardized residuals (standardization of the difference between observed and predicted values) are shown in the right panel of the rows. The distribution results symmetrical with respect to the 0, with a standard deviation lower than 2.5. The figure was made using MatLab 2019a.

**Figure 7.**
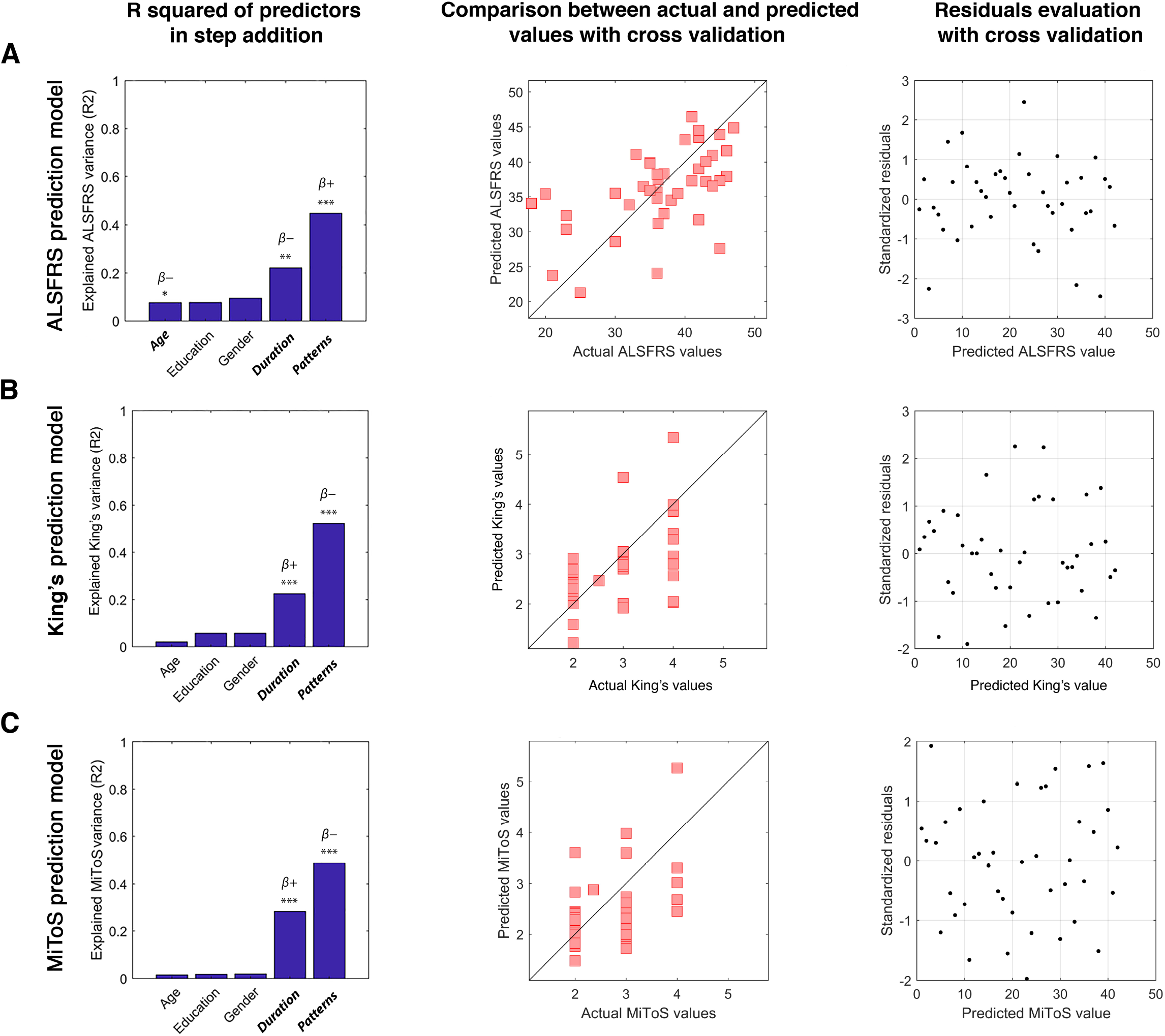
Multilinear model with *k*-fold cross-validation in the theta frequency band. Using as predictors age, education, gender, disease duration and number of patterns, the model predicts: (**A**) the ALSFRS-R (age: *P* = 0.034, *β* = -0.18; disease duration: *P* = 0.007, *β* = -0.05; number of patterns: *P* < 0.001, *β* = 0.05); (**B**) the King’s clinical staging system (disease duration: *P* = 0.002, *β* = 0.01; number of patterns: *P* < 0.001, *β* = -0.01); (**C**) the MiToS clinical staging system (disease duration: *P* < 0.001, *β* = 0.01; number of patterns: *P* = 0.002, *β* = -0.01). In the left panel of each row, the explained variance of the variable to be predicted as a function of the predictors is illustrated. Significant predictors are indicated in bold; positive and negative coefficients are illustrated with *β+* and *β-*, respectively; significance *P*-values: \**P* < 0.05, \*\**P* < 0.01, \*\*\**P* < 0.001. In the central panel of the rows, scatter plots of the comparison between actual and predicted values are represented. The standardized residuals (standardization of the difference between observed and predicted values) are shown in the right panel of the rows. The distribution results symmetrical with respect to the 0, with a standard deviation lower than 2.5. The figure was made using MatLab 2019a.

In the delta band the model provides significant predictions of the ALSFRS-R (**Fig. 6A**, *R*^*2*^ = 0.45), and both the King’s (**Fig. 6B**, *R*^*2*^ = 0.52) and the MiToS (**Fig. 6C**, *R*^*2*^ = 0.49) clinical staging systems. Similarly, in the theta band, the ALSFRS-R (**Fig. 7A**, *R*^*2*^ = 0.43), and both the King’s (**Fig. 7B**, *R*^*2*^ = 0.48) and the MiToS (**Fig. 7C**, *R*^*2*^ = 0.45) staging systems are predicted. The comparison between actual and predicted values and the residuals distribution obtained through the *k*-fold validation method^52^ are shown in the central and the right panel of each row of **Fig. 6** and **Fig. 7**, respectively. No significant contribution of education level and gender was observed.

We also validated our model using a LOOCV technique. Results were confirmed in both the delta (**Supplementary Fig. 3**) and the theta frequency bands (**Supplementary Fig. 4**).

## Discussion

In this work, we set out to predict clinical impairment in ALS in terms of efficient large-scale brain dynamics. Our results showed that ALS patients have a restricted functional repertoire as compared to healthy controls. This was demonstrated by the lower number of distinct (i.e., unique) avalanche patterns. The size of the functional repertoire correlates directly with the ALSFRS-R and negatively with both the King’s and the MiToS clinical staging systems.

It has been proposed that the healthy brain operates in a regime that maximizes flexibility of activations which, in turn, would underpin adaptive behaviour. With this in mind, we applied a recently developed framework to quantify the flexibility of fast brain dynamics^29^. Previous work showed that brain activity displays hyper-connected average topology of the functional brain networks in ALS^19,34^. The dynamic properties that lead to such average topological changes are not well understood. A working hypothesis might be that the brain in ALS is operating in a sub-optimal dynamical regime, resulting in more stereotyped activity. We borrow from statistical mechanics, a solid branch of physics, the concept of *neuronal avalanches*: these are fast, aperiodic bursts of activations that spread across the whole-brain^28^. If the brain is operating in a regime that allows flexible activity, neuronal avalanches efficiently reconfigure themselves over time^46^. All the unique patterns that have occurred (i.e. the states that have been visited by the brain) constitute the *functional repertoire*, and its size is used as a proxy for efficient fast brain dynamics^29^.

To sample the functional repertoire with sufficient spatio-temporal resolution, we used source-reconstructed resting-state MEG data acquired from a cohort of ALS patients and matched healthy controls.

We focused on the brain flexibility because it is now known that efficient reconfigurations of brain activated areas are linked to healthy brain functioning^53^. Therefore, the restriction in the number of avalanche patterns observed in ALS patients might reflect the effect of pathophysiological changes on the large-scale brain dynamics, as previously observed in other neurodegenerative diseases^29,54^.

Intriguingly, our results were specific to the delta and the theta frequency bands. However, alterations in regional power or static functional connectivity have been described in all frequency bands^55–58^. Ours is the first M/EEG study directly addressing the dynamic, aperiodic, scale-free activity in ALS. Hence, on the one hand, comparing our results with previous results, that were based on power-spectra or static connectivity, is not trivial. On the other hand, preliminary evidence from fMRI shows altered low-frequency brain dynamics in ALS^59^, which corroborates our results (given the comparable time-scales). However, given the different technique and type of analysis, comparing the results should be done cautiously. More dynamical M/EEG studies are warranted to confirm that the dynamical alterations in ALS are more prominent in slow time-scales.

In order to test whether some brain regions were specifically important in determining pathological patterns of activity, we analyzed shared and group-specific avalanche patterns. We classified avalanche patterns either as ‘shared’, if they occurred in both groups, or as ‘group-specific’, if they occurred in either group. Our results highlighted several brain regions being recruited significantly more often in ALS-specific neuronal patterns. In particular, basal ganglia were more often involved in avalanches in the ALS group than in the healthy controls, in both the delta and the theta frequency bands. This finding may support the key role of sub-cortical regions in recruiting cortical areas and supporting coherent activity across the brain^60^. The evidence also corroborates the wide-spread involvement of the brain in ALS^61^.

In fact, there is converging evidence showing marked atrophy in the hippocampus and in the basal ganglia in ALS. In particular, the thalamus, the putamen, the pallidum, the caudate and the nucleus accumbens show atrophy in ALS. Intriguingly, these regions are directly connected to cortical areas typically affected in ALS^62–64^. Furthermore, these sub-cortical structural modifications are closely associated with changes in cognitive and behavioral functioning in ALS^63,65,66^.

Subsequently, we reasoned that, if the *functional repertoire* is capturing a pathophysiological process, then it should be related to the stage of the disease and, in turn, allow its prediction. We found, as said, a positive correlation with the ALSFRS-R and a negative correlation with both the King’s and the MiToS clinical staging systems. Our findings might be compatible with the hypothesis that the neuropathological mechanisms in ALS shift the operational regime of the brain to a (presumably) sub-optimal state that no longer allows sufficient flexibility to support correct behavior, thereby relating to disability (as measured by the ALSFRS-R).

Consistent with our work, other time-resolved approaches to investigate functional connectivity have been used to link brain dynamics to disease severity. In particular, reduced temporal variability in network dynamics was found in ALS patients as compared to healthy controls. These changes significantly correlated with disease severity, in accordance with our results^67^.

The fact that the characterization of brain dynamics yields clinically relevant information regardless of the specific technique bears promise on the application of this framework to measure, non-invasively, pathophysiological processes in patients.

In this line of thinking, we investigated if brain dynamics could predict the ALSFRS-R and the stage of disease using a multilinear model analysis. Our results showed that the size of the functional repertoire is a significant predictor of both clinical staging and impairment, even after accounting for age, education, gender and disease duration. The ability of the functional repertoire to predict clinical disability suggests that brain flexibility is affected by the pathophysiological mechanisms and, consequently, could be utilized to measure them non-invasively.

Our work lays the foundation to study the effects that structural neurodegeneration and brain atrophy have on the spatio-temporal unfolding of the brain dynamics and, in turn, on behavioral outcomes.

## Conclusions

Our work shows that: 1) pathophysiological changes in ALS are reflected in reduced flexibility and, possibly, less effective communication; 2) sub-cortical regions contribute to brain dynamics and are affected by the pathophysiological processes of ALS. However, we found these regions from a post-hoc analysis that was not corrected for multiple comparison. Hence, this finding should be regarded as merely explorative; 3) the reduction of flexibility in ALS predicts disease stage as well as clinical impairment.

Based on these findings, we show that the framework of “neuronal avalanches” constitutes a straightforward, yet mathematically grounded, approach to non-invasively measure alterations in the brain dynamics. This, in turn, has potential for diagnostic applications as well as to lay hypothesis for targeted therapeutic approaches.

## Data Availability

All data produced in the present study are available upon reasonable request to the authors

## Abbreviations

AAL: Automated Anatomical Labeling
ALS: Amyotrophic Lateral Sclerosis
ALSFRS-R: Amyotrophic Lateral Sclerosis Functional Rating Scale-Revised
bvFTD: behavioral variant of Frontotemporal Dementia
FDR: False Discovery Rate
ICA: Independent Component Analysis
LCMV: Linearly Constrained Minimum Variance
LOOCV: leave-one-out cross validation
MEG: Magnetoencephalography
MiToS: Milano-Torino Staging
PCA: Principal Component Analysis
ROIs: Regions of Interest
TDP-43: TAR-DNA binding protein-43

## Supplementary material

Supplementary material is available at *Brain* online.

