## Supplementary materials for "Flexibility of brain dynamics predicts clinical impairment in Amyotrophic Lateral Sclerosis"

**Author affiliations:**

### Supplementary Figures

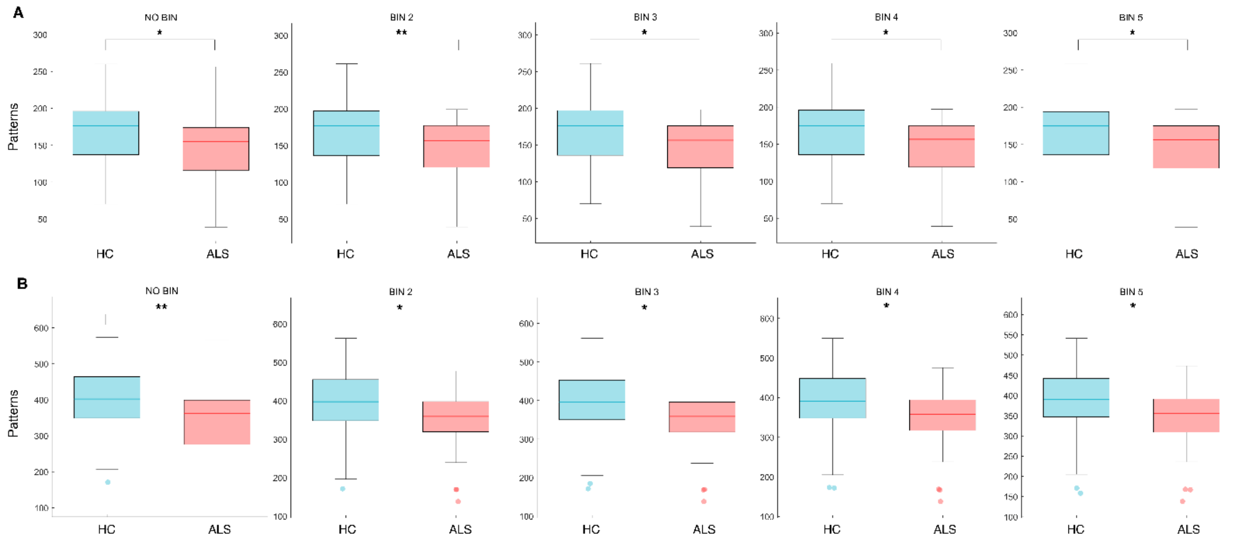


**Supplementary Figure 1. Comparison of the number of unique avalanche patterns.** (**A**) Box plots illustrating differences in the size of the functional repertoire in healthy controls (HC) and ALS patients (ALS) with different binnings in delta frequency band. For no binning, *P* = 0.010; for binning = 2, *P* = 0.009; for binning = 3, *P* = 0.010; for binning = 4, *P* = 0.010; for binning = 5, *P* = 0.011. (**B**) Box plots illustrating differences in the size of the functional repertoire in HC and ALS patients with different binnings in theta frequency band. For no binning, *P* = 0.006; for binning = 2, *P* = 0.010; for binning = 3, *P* = 0.012; for binning = 4, *P* = 0.015; for binning = 5, *P* = 0.019. Significance *P*-value: **P* < 0.05, ***P* < 0.01.

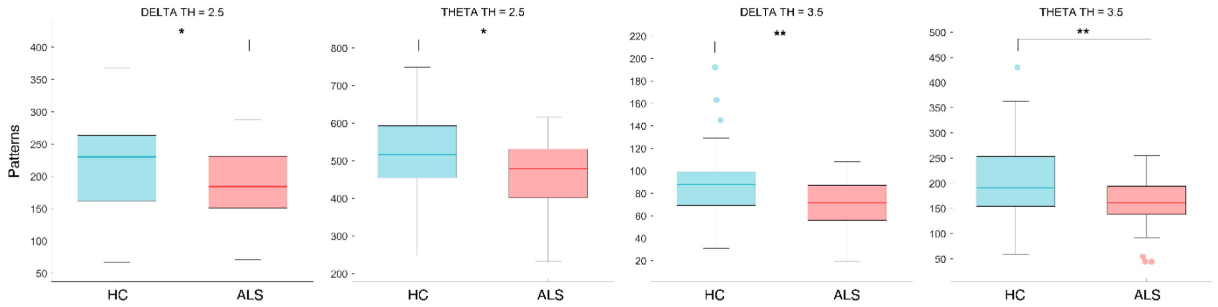


**Supplementary Figure 2. Comparison of the number of unique avalanche patterns.** Box plots illustrating differences in the size of the functional repertoire in healthy controls (HC) and ALS patients (ALS) with different thresholds. For $z$ = 2.5, $P$ = 0.028 in delta frequency band and $P$ = 0.030 in theta frequency band, while for $z$ = 3.5, $P$ = 0.005 and $P$ = 0.007 in delta and theta bands, respectively. Significance $P$-values: *$P$ < 0.05, **$P$ < 0.01.

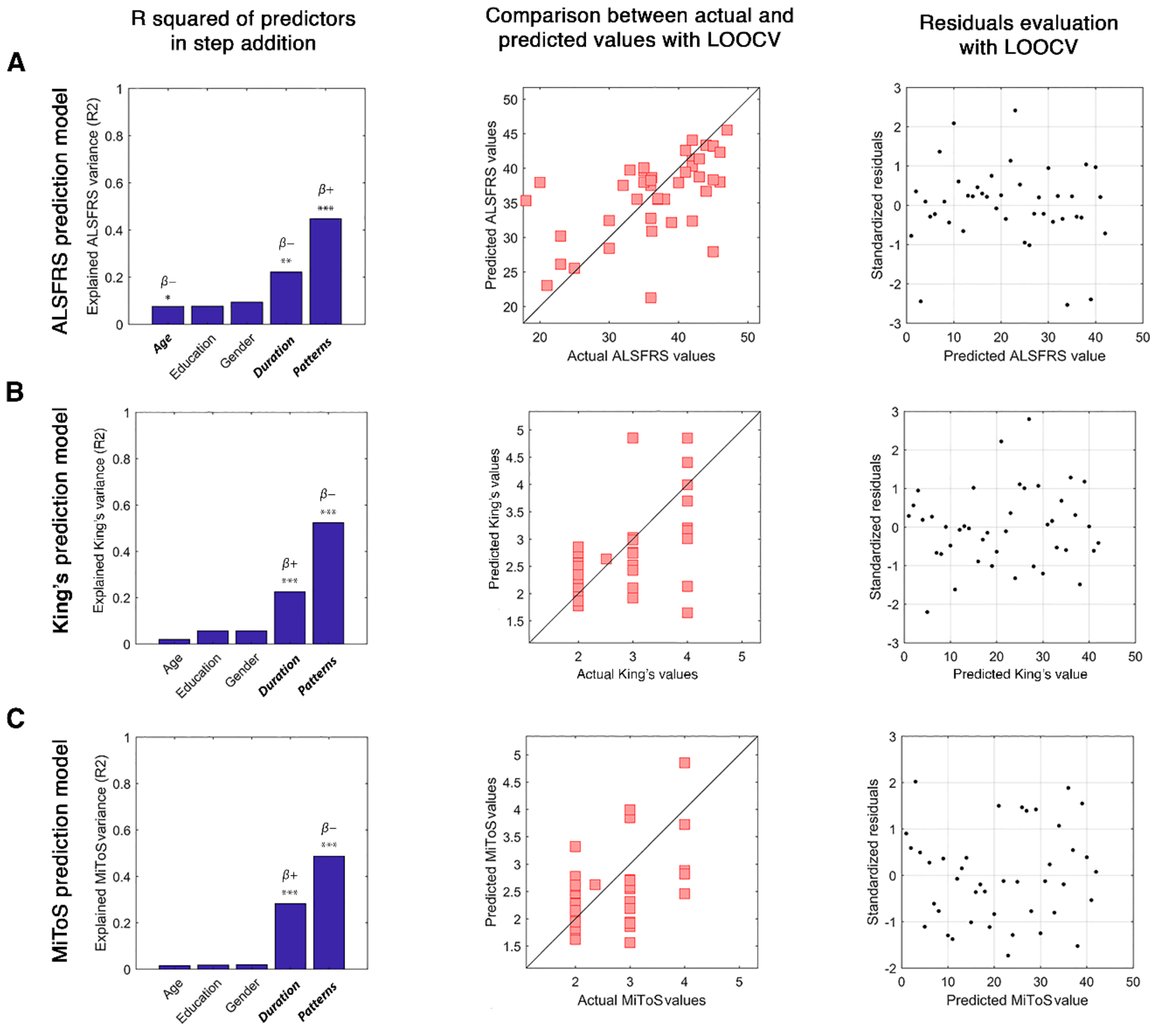


**Supplementary Figure 3. Multilinear model with leave-one-out cross-validation (LOOCV) in the delta frequency band.** Using as predictors age, education, gender, disease duration and number of patterns, the model predicts: (**A**) the ALSFRS-R (age: $P$ = 0.036, $\beta$ = -0.17; disease duration: $P$ = 0.004, $\beta$ = -0.06; number of patterns: $P$ < 0.001, $\beta$ = 0.11); (**B**) the King's clinical staging system (disease duration: $P$ < 0.001, $\beta$ = 0.01; number of patterns: $P$ < 0.001, $\beta$ = -0.02); (**C**) the MiToS clinical staging system (disease duration: $P$ < 0.001, $\beta$ = 0.01; number of patterns: $P$ < 0.001, $\beta$ = -0.01).

In the left panel of each row, the explained variance of the variable to be predicted as a function of the predictors is illustrated. Significant predictors are indicated in bold; positive and negative coefficients are illustrated with $\beta$+ and $\beta$-, respectively; significant $P$-values: *$P$ < 0.05, **$P$ < 0.01, ***$P$ < 0.001. In the central panel of the rows, scatter plots of the comparison between actual and predicted values with LOOCV are represented. The standardized residuals (standardization of the difference between observed and predicted values) are shown in the right panel of the rows. The distribution results symmetrical with respect to the 0, with a standard deviation lower than 2.5. The figure was made using MatLab 2019a.

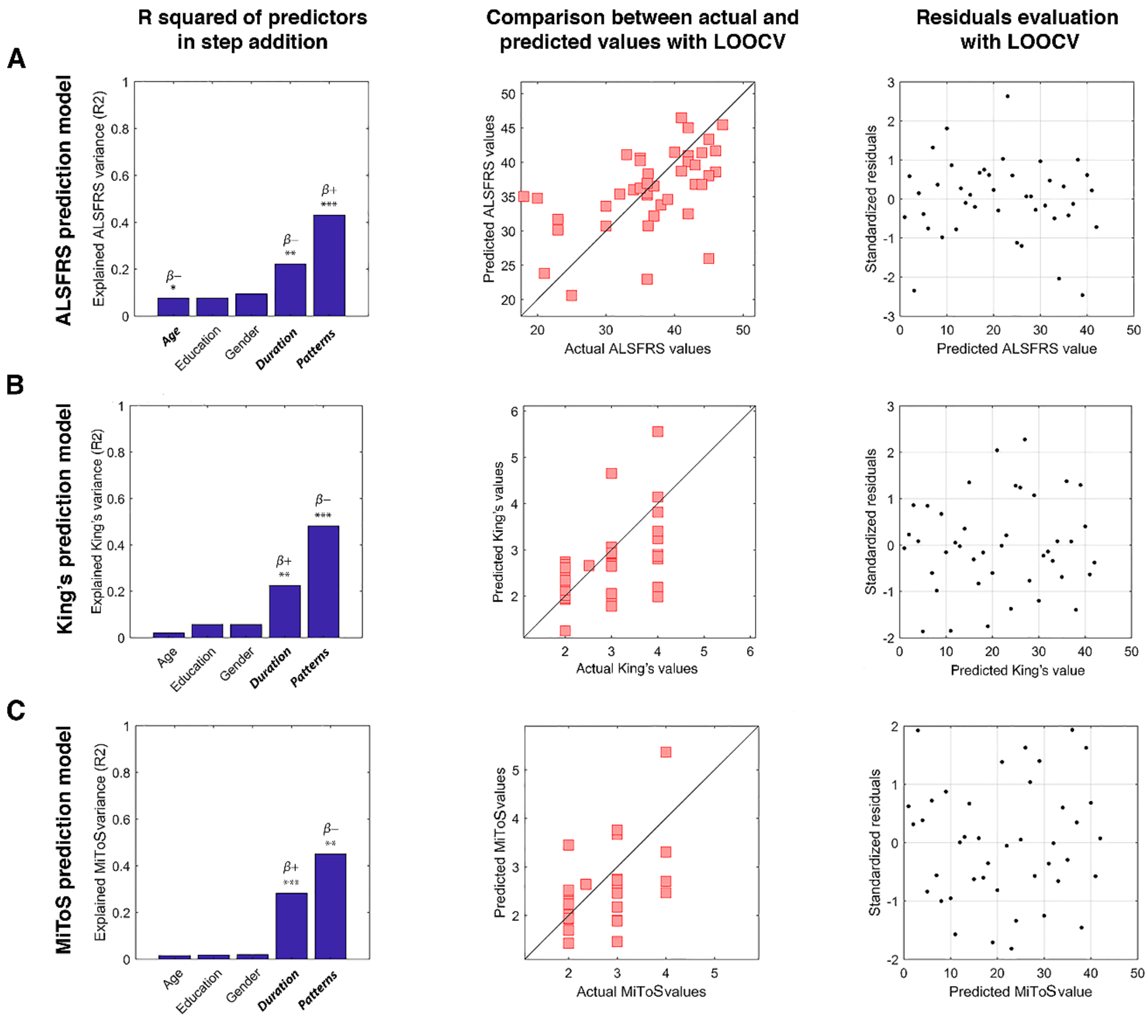


**Supplementary Figure 4. Multilinear model with leave-one-out cross-validation (LOOCV) in the theta frequency band.** Using as predictors age, education, gender, disease duration and number of patterns, the model predicts: (**A**) the ALSFRS-R (age: $P = 0.034$, $\beta = -0.18$; disease duration: $P = 0.007$, $\beta = -0.05$; number of patterns: $P < 0.001$, $\beta = 0.05$); (**B**) the King's clinical staging system (disease duration: $P = 0.002$, $\beta = 0.01$; number of patterns: $P < 0.001$, $\beta = -0.01$); (**C**) the MiToS clinical staging system (disease duration: $P < 0.001$, $\beta = 0.01$; number of patterns: $P = 0.002$, $\beta = -0.01$).

In the left panel of each row, the explained variance of the variable to be predicted as a function of the predictors is illustrated. Significant predictors are indicated in bold; positive and negative coefficients are illustrated with $\beta$+ and $\beta$-, respectively; significant *P*-values: *$P < 0.05$, **$P < 0.01$, ***$P < 0.001$. In the central panel of the rows, scatter plots of the comparison between actual and predicted values with LOOCV are represented. The standardized residuals (standardization of the difference between observed and predicted values) are shown in the right panel of the rows. The distribution results symmetrical with respect to the 0, with a standard deviation lower than 2.5. The figure was made using MatLab 2019a.
